# Multidimensional vulnerability and financial risk protection in health in contexts of protracted conflict: Evidence from the Occupied Palestinian Territory

**DOI:** 10.1101/2024.02.26.24303375

**Authors:** Julia Hatamyar, Sally Shayeb, Akseer Hussain, Weeam Hammoudeh, Sumit Mazumdar, Rodrigo Moreno-Serra

## Abstract

This paper proposes a multidimensional vulnerability index for a setting of protracted conflict, which is applied to study the relationship between financial vulnerability and catastrophic healthcare expenditure (CHE) incidence in the Occupied Palestinian Territory in 2018. We find that our index better captures the extent of financial risk protection (FRP) compared to conventional measures of financial welfare. Results indicate that the most vulnerable groups experience a significantly higher likelihood of incurring CHE, and this likelihood is increased for those living in the West Bank compared to the Gaza Strip. We also find a lack of protection from existing insurance types against the risk of CHE. Our analysis provides valuable insights about key aspects, such as health financing and insurance bottlenecks, that will deserve careful policy attention in efforts to rebuild the Palestinian health system, following the Israel-Hamas war.

**KEY MESSAGES:** *What is already known on this topic:*

• In settings of protracted conflict, conventional welfare measures, such as household consumption expenditure, may not adequately capture the multifaceted nature of financial risk protection (FRP) in health.
• There is a need for more comprehensive approaches to assess household vulnerability and FRP in such settings.

*What this study adds:*

• We propose a novel multidimensional index of household vulnerability for populations in protracted conflict areas, applied to 2018 data from the Occupied Palestinian Territory.
• Assessing FRP through this multidimensional lens reveals different patterns of exposure to financially catastrophic health expenditure (CHE) across sub-populations, which are not evident through traditional measures.
• We find a positive association between CHE risk and greater vulnerability in both the West Bank and the Gaza Strip, with the most vulnerable groups likely to incur CHE regardless of insurance status.

*How this study might affect research, practice, or policy:*

• Our vulnerability index predicts the risk of CHE across population sub-groups in a protracted conflict setting more effectively than traditional metrics, thereby offering better insights for health policy.
• The analysis highlights particular policy aspects, such as health insurance arrangements, that will require addressing to “build back better” the Palestinian heath system following escalation of violent conflicts, damages caused to critical health service and social infrastructure, and different constraints on available policy options.

## 1. INTRODUCTION

In settings with protracted conflict and other humanitarian challenges, conceptualisation of household vulnerability and its association with financial risk protection (FRP) in health may require a broader perspective than in more stable and peaceful areas. Conventional welfare measures such as household income or consumption expenditure, generally used in health financing research, may not fully capture the multi-faceted sources and aspects of vulnerability present in these unique settings, where threat of violence, political turmoil, forced displacement and movement restrictions can play a large role in individual well-being and financial risks. In such settings, measures of economic capacities need to be supplemented with appropriate, objective indicators from other contextually-relevant dimensions of vulnerability. Indicators that capture insecurities caused by the conflict in key areas of human security and welfare – food, water, shelter, freedom of movement, risks of injury, serious trauma and disabilities – are crucial to conceptualisations of vulnerability and realistic assessments of FRP. Measures of FRP using conventional income- or expenditure-centric measures of welfare, as we aim to illustrate, may significantly underestimate the levels and patterns of catastrophic expenditure or financial impoverishment due to high household spending on health care.

In this paper, we develop a multidimensional index of household vulnerability for populations experiencing protracted conflict applied in the Occupied Palestinian Territory (OPT) and examine its relationship with catastrophic health expenditure. The OPT exhibits a complicated political and economic situation [1]. Divided across the West Bank, Gaza strip and East Jerusalem, the region has witnessed protracted conflict and associated humanitarian consequences since 1948 [2]. Population health has faced detrimental consequences arising from extended periods of colonisation, along with other challenges, notably the fragmentation of the Palestinian health system and a reliance on international aid [3–5].

Using our multidimensional index, we observe how financial risk protection varies between the two major regions of the OPT – the West Bank (WB) and the Gaza Strip (GS) – and across other stratifying characteristics (e.g. sub-regions within West Bank, type of locality, education/occupation). We also aim to contribute to literature seeking to adjust FRP measurement in contexts with low access to healthcare services. In areas such as Gaza, which are more heavily affected by conflict violence and have even poorer health care infrastructure, we observe how such restricted access affects the demand for health care, consequently depressing health care expenditure. We identify several questions for future research, paving the way for deeper investigation into the complexities of healthcare access and FRP in challenging contexts.

**FIGURE 1.**
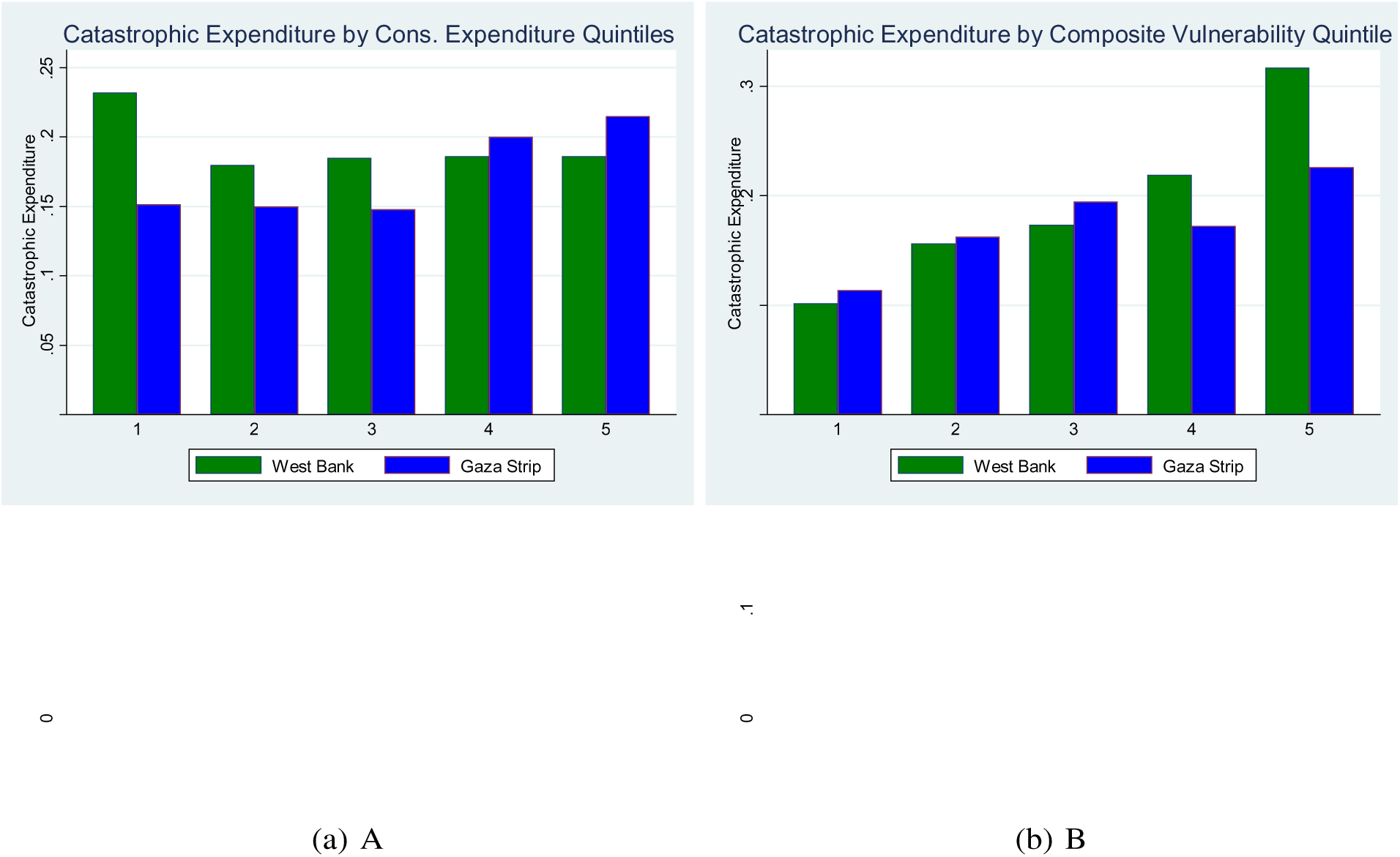
Catastrophic Health Expenditure Comparisons

Using data from the 2018 Socio-Economic Conditions (SEFSec) Survey, Figure 1 visually depicts the motivation for this work and shows that a conventional determinant of catastrophic health expenditure, overall consumption expenditure, may not accurately assess risk of CHE in the context of Palestine. There is an inverse relationship across the two regions of Palestine (WB and GS) between consumption expenditure and catastrophic health expenditure (Panel A). In WB, those with higher consumption expenditure are less likely to experience CHE, while in Gaza, the opposite holds true. Conversely, our composite vulnerability measure (described in Section 4) is consistently rising with catastrophic expenditure in both regions (Panel B).

We begin with a brief overview of the Palestinian context and a review of the literature on financial vulnerability in conflict affected areas. In the following sections, we show that our composite vulnerability measure is capable of effectively capturing information about the likelihood of incurring CHE, and explore the heterogeneous effects of vulnerability across regions, insurance type and status, and locality (urban versus rural dwelling, refugee camps).

### 1.1 Background and Motivation

#### Conflict Affected Areas and Health Services

Conflict, whether internal or international, is a crucial social determinant of health outcomes [6], directly and indirectly jeopardising access to essential services [7–9], appropriate nutrition, and healthcare facilities [10–13]. Conflict often does not end with clear outcomes like victory or peace agreement; instead, it frequently leads to a state of limbo with fighting simply ceasing [14]. The transition to a post-conflict situation is hardly ever linear, and some nations can relapse into conflict [15, 16]. The protracted conflict has immediate consequences, but also longterm impacts influencing multiple generations. It can affect economic access, social stability, healthcare availability, and access to health facilities [17–19].

Conflict-affected states typically experience worse health service provision [15, 20, 21] compared to non-conflict-affected states [22]. This situation is exacerbated by factors like high out-of-pocket payments, low government health expenditure, and external dependency on funding [23]. Conflict-affected states encounter various challenges in delivering healthcare, including ideological and operational conflicts among service providers [24, 25] and issues related to care for internally displaced persons and refugees in countries with established health systems [26]. Improving healthcare provision in these regions can be achieved through public financing, efficient utilisation of resources by governments [23], performance-based financing [27], subsidies [28], and community involvement [29]. These measures are essential to advance universal health coverage and address health disparities prevalent in these challenging environments.

#### Overview of the Palestinian Conflict and Health System

The formation and structure of the Palestinian health system is deeply rooted in its complex political history. The OPT have been in political turmoil and under protracted conflict for many decades, with implications for public services such as health, education and social protection [4]. Since the Oslo Agreement of 1993, which aimed to achieve peace between the Israeli government and the Palestine Liberation Organisation (PLO) and led to the establishment of the Palestinian Authority (PA), there has been noticeable decline in Palestinian economic standards, worsening social degradation, and an acceleration of the de-development process [2].^1^ These issues are compounded by unenforced international human rights legislation, internal Palestinian divides, and confrontations between Israeli and Palestinian armed organisations on a regular basis [30]. One of the outcomes of the protracted conflict between the governments of Israel and Palestine is enclavisation, as witnessed in the physical separation of the West Bank and the Gaza strip [2, 31]. The resulting absence of a long-term, unified political settlement and prospects for future development weakened an already frail public-sector structure, particularly in health-care delivery [30].

From 1967, following the occupation of the OPT and until 1995, the Israeli Civil Administration managed the governmental health care system, which was consistently underfunded and limited in terms of health and social services coverage [4, 32]. Severe budget constraints, referral to Israeli hospitals for tertiary care, and restrictions on licenses for new healthcare projects resulted in the OPT’s total reliance on the Israeli health system [1]. Following the Oslo Accords, health governance responsibilities were partially transferred to the newly established Palestinian Authority (PA) in 1994, which attempted to build a more comprehensively structured health system and established the Ministry of Health [33]. Between 1994 and the onset of the Second Intifada in 2000, efforts were made to standardise health services, improve health infrastructure, and train healthcare professionals. International donor assistance increased during this brief period, allowing for further development of healthcare infrastructure, though new challenges were introduced by inconsistency in funding and complex coordination efforts required to distribute aid. The renewal of conflict during the Second Intifada (2000-2005) severely impacted the nascent healthcare system, causing damage to the new infrastructure, restricting movement and thereby access to healthcare, increasing the medical needs of the population [34]. Local systems were unable to handle these needs, increasing reliance on referrals to outside healthcare providers [35].

Tensions escalated between Hamas, the Islamic resistance movement which won a majority of seats in the 2006 Palestinian Legislative Council elections, and Fatah, the Palestinian national liberation movement, which had largely controlled the West Bank’s PA, leading to several violent clashes and culminating in the seizure of the Gaza strip by Hamas in 2007. Since then, the West Bank and Gaza have been under separate political leaderships, with the PA having limited governance in the West Bank and Hamas with limited control in the Gaza Strip under prolonged blockade. Following Hamas’ takeover, Israel began blockading the Gaza strip with land, sea, and air closures.

The blockade has severely limited the movement of people and goods in and out of the area – including healthcare professionals, patients in need of complex procedures unavailable within Gaza, and health supplies [4, 36, 37]. The geopolitical split has led to a fragmented health system between the two regions, with challenges in governance, resource allocation, and health strategy coordination. Decreased tax revenue and foreign aid created financial instability, worsening the ability of the PA to maintain healthcare services [5, 38]. These conditions comprised the baseline of health and social services prior to the Israel-Gaza war beginning in October 2023. It is worth noting that the degree and magnitude of destruction of infrastructure, as well as deaths, injuries, and displacement from Israeli attacks on the Gaza Strip will have grave repercussions for the health system. As of 12 February 2024, this has amounted to: 28,340 Palestinians killed in the Gaza Strip and 67,984 Palestinians injured, including at least 12,300 children (UN-Office for the Coordination of Humanitarian Affairs^2^).

Despite PA efforts toward establishing adequate social protection [39, 40], there is an anticipated rise in the rate of poverty across the life cycle in the OPT [39]. Governmental and non-governmental assistance vary in coverage and impact. Additionally, vulnerable groups such as individuals with disabilities are not adequately addressed by existing social protection policies [39]. Furthermore, health inequalities are the result of a complex interplay of variables including economic difficulties, food shortages, environmental exposures, psychological trauma and stress, and limited access to health care. Most of these factors may be directly or indirectly linked to Israel’s military occupation of the OPT [41]. Indeed, frequency of attacks on medical missions is correlated with peaks in occupation-related injuries and fatalities [42].

#### Conceptualising Vulnerability in Protracted Conflict Settings: Health Financing, FRP, and CHE

The assessment of socioeconomic inequities encountered by disadvantaged communities is especially important in protracted conflict settings. Previous work has shown that creation of multidimension.al indices for use in the study of health expenditure and other outcomes can provide benefits beyond use of individual measures alone.

In making these assessments. Pinilla-Roncancio, Amaya-Lara, Cedeño-Ocampo, Rodriguez-Lesmes, and Sepúlveda [43] create a multidimensional poverty index using a modified Alkire-Foster technique [44], capturing information such as access to clean water, sanitation facilities and electricity, as well as aspects related to mitigation of negative health shocks, such as asset ownership and employment status. The authors then examine its relationship with CHE in seven different LMICs - importantly, each country requires its own unique index depending on heterogeneous characteristics and data availability - and show that CHE incidence can drive long term poverty. In Palestine, such an index has been used by the Bureau of Statistics and included both monetary and non-monetary elements, to expand the scope of poverty studied, but without connecting the index to health expenditures (although information such as disability prevalence, presence of chronic diseases, health insurance and health facility accessibility are used in creation of the index) [45].

Without basic human security, international aid directed towards provision of healthcare in Palestine is unable to promote its development [36]. Vulnerability to shocks of a more fundamental and physical nature than financial poverty is more likely to impact risk protection in health in this context. Direct threats to security faced by Palestinians, such as restrictions on movement, arrests and detentions, gunfire, home demolition, injury and death, will not be captured in a poverty index. Survey data (see Section 3) show the collective history of mass displacement and being repeatedly uprooted have resulted in commonly experienced feelings of insecurity and instability. Therefore, in this paper we develop a composite vulnerability index that aims to capture complex dynamics of CHE, which is often related to poverty but may encompass other aspects in settings of protracted conflict. Examining CHE through this index can assist in measuring household financial resilience and coping methods in the face of healthcare payments, better reflecting the degree of FRP - or lack thereof - among the most vulnerable groups in a long-term conflict scenario.

## 2. METHODS

### 2.1 Data and Variables

In this study the empirical analyses are based on data from the 2018 Socio-Economic Conditions Survey conducted by the Palestinian Central Bureau of Statistics (PCBS).^3^ SEFSec-2018 is the third round of the panel survey, which includes respondents from 9926 families.

The outcome variables of interest in our analysis are catastrophic health expenditures (CHE) as a percentage of household consumption expenditure (at the 10% threshold), and household non-food expenditure (at the 20% threshold). The 10% threshold is commonly used by global organisations to assess FRP and monitor progress towards UHC [46]. The main variable of interest that we use to examine variation in the CHE outcome is the *vulnerability index*. To create the vulnerability index, a factor analysis was performed using a number of survey questions capturing both objective and subjective measures. The factor analysis is done separately for each region, West Bank and Gaza (as factor loadings may be different, given the significant regional differences shown in Table 1), and a composite vulnerability score is predicted from these factors. Using the principal factor method in Stata, factors are retained if their eigenvalues of the covariance matrix exceed the value of 1. Our vulnerability index was finally obtained by assigning scores to each observation. These scores are weighted averages of the covariates, with weights determined by the factor loadings (coefficients indicating the strength of the association between each variable and the factor).

Our vulnerability index is similar in spirit to the concept of human insecurity explored in Palestine by Ziadni et al. [37], who describe insecurity comprising two aspects: basic material needs required for survival, and a psychological/social component [47]. Factors used in our vulnerability index composition are: self-assessed economic status, self-assessed household resilience (whether a household is able to keep up financially), self-assessed need for public (financial) assistance, and a combined shocks experience variable which includes political, economic, health, educational, freedom, water availability, and food-related shocks, asset ownership (calculated using principal component analysis), self-assessed deprivation level, and a human insecurity scale. Appendix Table A2 shows the vulnerability index factor loadings for both regions, and indicates differences in loadings between the two regions. For the main analyses terciles of vulnerability composite index are used.^4^ For multivariate analyses, we include relevant covariates, such as employment status categories, education level of household head, presence of disability or other chronic health conditions, health insurance status and type; and number of household members.^5^

**TABLE 1.** Summary Statistics By Region.

| **Socioeconomic Variables** | WB<br>N=5895 | Gaza<br>N=4025 | Mean Diff | p-value |
| --- | --- | --- | --- | --- |
| Not Working | 0.264 | 0.496 | -0.23*** | 0.000 |
| Part-Time Work | 0.103 | 0.128 | -0.02*** | 0.000 |
| Full-Time Work | 0.338 | 0.211 | 0.13*** | 0.000 |
| Long Work Hours | 0.296 | 0.165 | 0.13*** | 0.000 |
| Elementary or Less Education | 0.319 | 0.253 | 0.07*** | 0.000 |
| Preparatory Education | 0.326 | 0.279 | 0.05*** | 0.000 |
| Secondary Education | 0.160 | 0.183 | -0.02** | 0.004 |
| Above Secondary Education | 0.195 | 0.286 | -0.09*** | 0.000 |
| No Disability/Chronic Condition | 0.715 | 0.638 | 0.08*** | 0.000 |
| Chronic Condition Only | 0.143 | 0.119 | 0.02*** | 0.000 |
| Disability Only | 0.061 | 0.141 | -0.08*** | 0.000 |
| Chronic and Disability | 0.081 | 0.102 | -0.02*** | 0.000 |
| No Insurance | 0.299 | 0.064 | 0.23*** | 0.000 |
| PA only | 0.379 | 0.270 | 0.11*** | 0.000 |
| UNRWA only | 0.126 | 0.162 | -0.04*** | 0.000 |
| PA+UNRWA | 0.116 | 0.493 | -0.38*** | 0.000 |
| Urban | 0.692 | 0.847 | -0.16*** | 0.000 |
| Rural | 0.243 | 0.000 | 0.24*** | 0.000 |
| Camps | 0.065 | 0.153 | -0.09*** | 0.000 |
| HH members | 4.737 | 5.621 | -0.88*** | 0.000 |
| Dependency Ratio | 0.740 | 0.881 | -0.14*** | 0.000 |
| Healthcare Facility Access Difficulty | 0.193 | 0.270 | -0.08*** | 0.000 |
| Received Any Assistance | 0.091 | 0.676 | -0.59*** | 0.000 |
| Refugee | 0.266 | 0.615 | -0.35*** | 0.000 |
| **Vulnerability Index Components** | WB | Gaza | Mean Diff | p-value |
| Self-Assessed Poverty | 0.103 | 0.478 | -0.37*** | 0.000 |
| Self-Assessed Financial Fragility | 0.207 | 0.643 | -0.44*** | 0.000 |
| Subjective Need for Assistance | 0.442 | 1.456 | -1.01*** | 0.000 |
| Political Conflict Shock | 0.122 | 0.042 | 0.08*** | 0.000 |
| Economic Shock | 0.203 | 0.610 | -0.41*** | 0.000 |
| Health/Education Shock | 0.131 | 0.541 | -0.41*** | 0.000 |
| Freedom Shock | 0.027 | 0.102 | -0.07*** | 0.000 |
| Water Shock | 0.323 | 0.537 | -0.21*** | 0.000 |
| Food Security Shock | 0.196 | 0.797 | -0.60*** | 0.000 |
| Asset Ownership Index | 2.364 | 1.600 | 0.76*** | 0.000 |
| Subjective Deprivation | 1.437 | 2.666 | -1.23*** | 0.000 |
| Human Insecurity Scale | 36.006 | 41.422 | -5.42*** | 0.000 |
| **CHE Measures** | WB | Gaza | Mean Diff | p-value |
| 10% Consumption Exp. | 0.352 | 0.274 | 0.08*** | 0.000 |
| 20% Non-food Exp. | 0.134 | 0.103 | 0.03*** | 0.000 |

Table 1 shows mean-differences and p-values by region for all covariates. The two regions of the OPT differ immensely in terms of healthcare, educational, and occupational characteristics, likely due to the closure of nearly 80% of industry in Gaza after the Israeli blockade [37]. Compared to West Bank residents, individuals living in Gaza are more likely to be unemployed, have more disabilities and chronic conditions, and live in urban areas and refugee camps. Gazans are much more likely to self-assess themselves as fragile, poor, and in need of financial assistance. They face larger economic and food security shocks by orders of magnitude compared to West Bank residents, who face larger political conflict shocks.

We also present information on incidence of CHE at various thresholds ranging from 5% to 40% of consumption and non-food expenditure and of total household expenditure by region in Appendix Table A1.^6^ We note that, regardless of threshold or definition of living standard/capacity to pay, CHE incidence is higher in the West Bank than in Gaza. Figure 2 visualises the intensity of the incidence of CHE at governorate levels. It demonstrates clear geographic variation in incidence, with southern areas of the West Bank experiencing more CHE than northern West Bank. For example, in Hebron, located in the Southern WB, more than 25% of respondents have faced CHE at the 10% threshold compared to the northern governorates of the West Bank.

**FIGURE 2.**
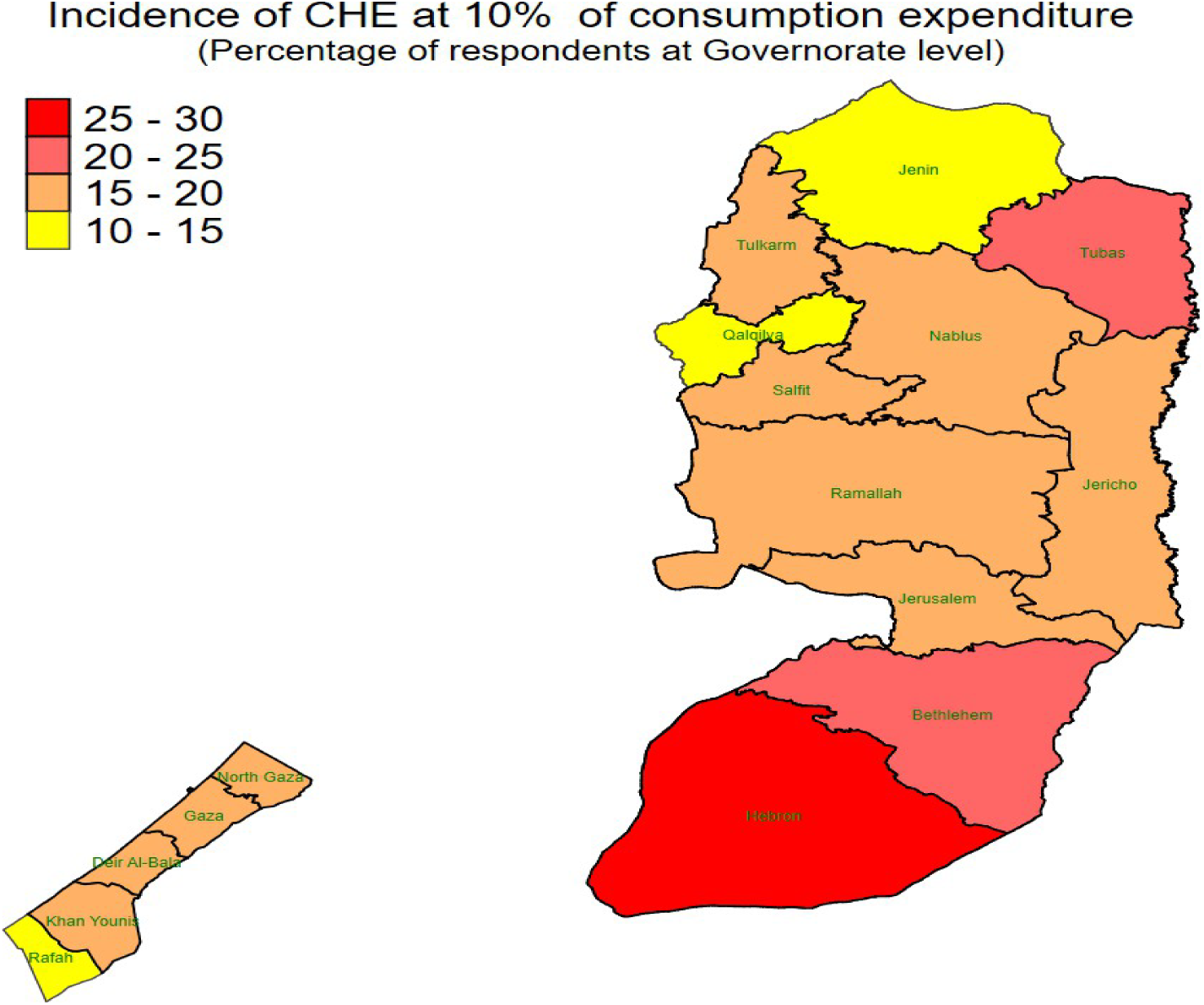
CHE by Governorate

### 2.2 Regression Model

For analysing the relative risks of experiencing CHE across different levels of multidimensional vulnerability, we run the following logistic regression:

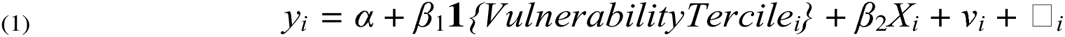

where *y_i_* is catastrophic health expenditure at the 10% threshold, **1***{VulnerabilityTercile}* is an indicator for being in a tercile of the vulnerability index, ν*_i_* is the governorate fixed effect, and _J*_i_* is the error term. Our suite of control covariates used in the primary analysis, *X_i_*, includes employment status, household size, insurance status and type, education level, and a variable indicating whether there is chronic illness or disability present in the household. Detailed descriptions of covariates can be found in the Appendix.

## 3. RESULTS

Here we present the main results of this paper, which show the association between multi-dimensional vulnerability and the likelihood of incurring CHE. Our results are robust across specifications, with the most vulnerable groups being significantly more likely to experience catastrophic health expenditures. The likelihood is higher in WB compared to Gaza.

As indicated in Table 2 Panel A, Palestinian households within the second and third terciles of the vulnerability index are significantly more likely to incur CHE at 10% of total expenditure and at 20% of non-food expenditure. The most vulnerable households are 76.2% and 105.3% more likely to incur CHE at the mentioned thresholds, respectively. In both specifications, WB experiences a higher likelihood of experiencing CHE in the most vulnerable tercile. Panel B of Table 2 compares the impact of quintiles of monthly percapita consumption expenditure (MPCE) with the impact of our vulnerability index on CHE at the 10% level of total expenditure. It is clear that the MPCE measure does not have a significant impact, nor any discernable pattern. For example, there is a slight impact of the fourth MPCE quintile in GS (OR 1.51), but the magnitude decreases for the highest quintile and does not remain significant. In contrast, our vulnerability largely shows a consistent increasing relationship with CHE. For the OPT as a whole and for the West Bank, the magnitude of coefficients is increasing with quintiles, and for GS the highest quintile also has the largest effect. We describe the effects of other (control) variables, which are often highly significant, in the Appendix.

**TABLE 2.** Catastrophic Health Expenditure and Vulnerability.

| **PANEL A: Consumption Expenditure vs. Non-food Expenditure** |  |  |  |  |  |  |
| --- | --- | --- | --- | --- | --- | --- |
| Dep Var: | CHE-10% |  |  | Non-food- 20% |  |  |
|  | (1) | (2) | (3) | (4) | (5) | (6) |
| Odd Ratios: | All | WB | Gaza | All | WB | Gaza |
| **Index Tercile = 2** | 1.264**<br>(0.141) | 1.135<br>(0.171) | 1.500**<br>(0.251) | 1.455***<br>(0.141) | 1.420**<br>(0.234) | 1.552***<br>(0.127) |
| **Index Tercile = 3** | 1.762***<br>(0.144) | 1.846***<br>(0.216) | 1.617***<br>(0.148) | 2.053***<br>(0.241) | 2.085***<br>(0.255) | 1.920***<br>(0.447) |
| *Observations* | 9647 | 5801 | 3846 | 9646 | 5800 | 3846 |
| *Clusters – Governorate* | 16 | 11 | 5 | 16 | 11 | 5 |
| **PANEL B: Consumption Expenditure vs. Vulnerability Index** |  |  |  |  |  |  |
| Quintile Definition: | MPCE |  |  | Vulnerability Index |  |  |
|  | (1) | (2) | (3) | (4) | (5) | (6) |
| Dep: Var: CHE-10% | All | WB | Ghaza | All | WB | Gaza |
| **Index Quintile=2** | 0.917<br>(0.089) | 0.834*<br>(0.086) | 1.088<br>(0.173) | 1.448***<br>(0.181) | 1.447**<br>(0.272) | 1.463*<br>(0.289) |
| **Index Quintile=3** | 0.958<br>(0.095) | 0.931<br>(0.101) | 1.058<br>(0.204) | 1.588***<br>(0.218) | 1.484<br>(0.379) | 1.782***<br>(0.201) |
| **Index Quintile=4** | 1.068<br>(0.143) | 0.893<br>(0.118) | 1.515*<br>(0.338) | 1.740***<br>(0.126) | 1.851***<br>(0.214) | 1.571***<br>(0.125) |
| **Index Quintile=5** | 0.994<br>(0.160) | 0.836<br>(0.115) | 1.499<br>(0.462) | 2.280***<br>(0.232) | 2.486***<br>(0.398) | 1.963***<br>(0.161) |
| *Observations* | 9887 | 5872 | 4015 | 9647 | 5801 | 3846 |
| *Clusters – Governorate* | 16 | 11 | 5 | 16 | 11 | 5 |

Panel A compares two different dependent variables: CHE at 10% total expenditure and 20% of non-food expenditure. Panel B compares two independent variables: monthly consumption expenditure versus the vulnerability index. Exponentiated coefficients are shown with standard errors in parentheses, clustered at governorate level. Governorate fixed effects in all models. All control covariate coefficients reported in Appendix. \* $p < 0.10$ , \*\* $p < 0.05$ , \*\*\* $p < 0.01$

### 3.1 Heterogeneity Analysis

#### CHE by Insurance Type

Due to initial findings of a highly significant and large relationship between governmental health insurance and likelihood of incurring CHE (see regression coefficients in Appendix), here we subset the data and repeat the analysis. First, we subset the data by insurance type, and then by insurance status. Results for regressions on data subsetted by insurance type and status are shown in Table 3.^7^ For those individuals only with governmental (PA) insurance, we see that the impact of being in the highest vulnerability tercile is slightly larger than that of the entire sample (1.78 in Panel A, versus 1.76 in Table A4), and the pattern is consistent across the two regions. Households with only governmental health insurance experience a statistically significant increase in the likelihood of CHE at 10% of total expenditure, signifying a 33.1% and 78.7% higher likelihood of incurring CHE in second and third vulnerability terciles (p*<*0.01). For Gaza, results show 87.6% and 58.3% higher likelihood of incurring CHE among second and third vulnerability terciles, respectively. Also of note is the lack of any association of the vulnerability terciles with CHE for those individuals only possessing UNRWA insurance. This suggests that the impact of insurance type on CHE varies across vulnerability levels, with a stronger effect for those in the higher vulnerability category who are not in possession of UNRWA insurance.

**TABLE 3.** Catastrophic Health Expenditures, Heterogeneity Analysis.

| **PANEL A: INSURANCE TYPE** |  |  |  |  |  |  |
| --- | --- | --- | --- | --- | --- | --- |
| Insurance Type: | PA Only |  |  | UNRWA Only |  |  |
|  | (1) | (2) | (3) | (4) | (5) | (6) |
| Dep: Var: CHE-10% | All | WB | Ghaza | All | WB | Gaza |
| **index tercile=2** | 1.331**<br>(0.178) | 1.133<br>(0.153) | 1.876***<br>(0.398) | 1.122<br>(0.229) | 1.225<br>(0.343) | 1.109<br>(0.409) |
| **index tercile=3** | 1.787***<br>(0.172) | 1.877***<br>(0.250) | 1.583***<br>(0.219) | 1.523<br>(0.402) | 1.481<br>(0.519) | 1.623<br>(0.752) |
| *Observations* | 3246 | 2212 | 1034 | 1352 | 728 | 624 |
| **PANEL B: Insurance Status** |  |  |  |  |  |  |
| Insurance Status: | Insured |  |  | Uninsured |  |  |
|  | (1) | (2) | (3) | (4) | (5) | (6) |
| Dep: Var: CHE-10% | All | WB | Ghaza | All | WB | Gaza |
| **index tercile=2** | 1.334**<br>(0.172) | 1.121<br>(0.168) | 1.620***<br>(0.300) | 1.337*<br>(0.226) | 1.337<br>(0.236) | 1.312<br>(1.010) |
| **index tercile=3** | 1.825***<br>(0.172) | 1.820***<br>(0.307) | 1.767***<br>(0.157) | 2.622***<br>(0.356) | 2.571***<br>(0.343) | 2.339<br>(2.540) |
| *Observations* | 7670 | 4063 | 3607 | 1972 | 1736 | 236 |
| **PANEL C: Locality Type** |  |  |  |  |  |  |
| Insurance Status: | Urban |  |  | Rural | Camps |  |
|  | (1) | (2) | (3) | (4) | (5) |  |
| Dep: Var: CHE-10% | All | WB | Gaza | All | All |  |
| **index tercile=2** | 1.403***<br>(0.157) | 1.212<br>(0.161) | 1.740***<br>(0.271) | 0.987<br>(0.245) | 1.115<br>(0.179) |  |
| **index tercile=3** | 1.966***<br>(0.112) | 1.979***<br>(0.184) | 1.987***<br>(0.136) | 2.077**<br>(0.591) | 1.648**<br>(0.384) |  |
| *Observations* | 7253 | 4008 | 3245 | 1417 | 972 |  |

Exponentiated coefficients; Standard errors in parentheses
SE clustered at governorate level, with governorate fixed effects in all models
\* $p < 0.10$ , \*\* $p < 0.05$ , \*\*\* $p < 0.01$

#### CHE by Insurance Status

In Panel B of Table 3, we report for data subsetted by insurance status. We find that those who have any type of insurance are less likely than the uninsured to incur CHE. Further, likelihood of incurring CHE increases with vulnerability index terciles, by 33.4% and 82.5% for the second and third terciles among insured households and 33.7% and 162% among uninsured households. The most vulnerable groups are more likely to incur CHE, regardless of health insurance status. Examining results by region, we find in WB, CHE risk increases by 82% among insured households (Panel B) compared to 87.7% among households with governmental health insurance only (Panel A). However, in the Gaza Strip, the most vulnerable tercile with *any* type of insurance is more likely to incur CHE than those with the PA type only (76% vs 58%).

In summary, the health insurance heterogeneity analysis suggests that association between health insurance type and the likelihood of incurring CHE is not uniform across vulnerability index terciles. The most vulnerable groups are more likely to incur CHE, regardless of their health insurance status. However, the type of health insurance may play a role in the likelihood of CHE for the most vulnerable groups.

#### CHE by Locality Type

We also compare results for data subsetted by locality type (urban, rural, and refugee camps). Results are shown in Table 3, Panel C. Results are consistent with the overall findings and show an increase in the likelihood of CHE as vulnerability increases, but also show that those living urban areas experience a greater likelihood of incurring CHE at the 10% threshold compared to the overall sample (40.3% for the second tercile, and 96.6% for the most vulnerable tercile in urban areas). For households residing in rural areas, the third tercile demonstrates a noteworthy OR of 2.077.

When comparing the most vulnerable living in urban areas across WB and GS, we see the difference between likelihood of incurring CHE at the 10% level is roughly equal. This indicates that households living in rural areas in WB may be driving the overall (full sample) results, as they are more likely to incur CHE than their urban counterparts. Additionally, population in GS is predominantly confined to densely populated urban and camp areas. Among households in refugee camps, the third tercile experiences a significant OR of 1.648.

### 3.2 Index Validity

Finally, as survey data contains subjective reporting of household well-being, we check whether the inclusion of self-assessed survey variables impacts the findings; results are depicted in Appendix Table A11. We recreated the vulnerability index by removing any subjective factors. This definition also ensured that there are no missing households for the vulnerability index, as around 250 households did not complete the supplementary mental health survey module. The main results of our paper were largely unchanged, indicating that these subjective survey questions or missing households were not driving the findings.

## 4. DISCUSSION

In this paper, we have proposed a multidimensional vulnerability index to study the relationship between financial vulnerability and catastrophic healthcare expenditure (CHE) incidence in the Occupied Palestinian Territory. We have found that the most vulnerable groups are more likely to incur CHE. The consistent relationship between vulnerability and CHE implies that the index captures vulnerability according to the Palestinian “experience”, irrespective of region, thus aiding in potential policy actions.

The finding of this paper that households in the West Bank are more likely to experience CHE compared to those in the Gaza Strip is likely related to healthcare utilisation and access differences across the two regions. The impact of conflict intensity on maternal healthcare utilisation (as measured by antenatal care visits) is greater in the South West Bank compared to the Gaza strip, where antenatal care may be seen as non-necessary and stressful during times when travel is more difficult and/or dangerous [35]. Concentration of healthcare facilities is also lowest in the southern West Bank. Distance to primary (basic) healthcare services is shorter within the Gaza strip, whereas individuals in the Southern WB face checkpoints and movement restrictions more regularly. However, to access more complex and specialised procedures, Gazans usually must be moved to WB hospitals, which requires a special permit [35, 38]. Since Gazan households simply are not able to utilise these more expensive healthcare services, their risk of CHE incidence may be reduced - while at the same time, their risk of adverse health outcomes increases. Moreover, between 2006 and 2022, Gaza’s real GDP per-capita declined by (37%), and its share in the Palestinian economy decreased from (31%) to (17.4%). Households experience a high probability of poverty (65%), labour force dropout (41%), and unemployment (45%) in this context [48].

Our puzzling finding that the possession of governmental health insurance may lead to higher likelihood of incurring CHE bears further discussion here. The current governmental health insurance scheme faces notable challenges to provision of comprehensive coverage. The scheme’s revenues only cover a small fraction (≤ 10%), of overall health service costs, highlighting a significant funding gap. Moreover, the voluntary nature of the system results in a limited subscription base, constraining its capacity to provide widespread FRP. Exemptions granted to various groups lack a compensatory mechanism, placing additional strain on the financial sustainability of the health insurance fund. The dual role of the Ministry of Health, which manages both the health insurance scheme and delivery of basic health services, creates a potential conflict of interest, impacting the quality and accessibility of health services. Costly patient transfers, often lacking defined controls and influenced by political or community pressures, further contribute to financial challenges. Additionally, long-term internal political divisions and the decision to exempt Gazans from health insurance fees have undermined expenditure control capabilities and deprive the health system of crucial revenues [49].

## 5. CONCLUSIONS

This work has uncovered significant financial vulnerability in health among population sub-groups in the OPT. We must emphasise, however, that our data analysis represents a snapshot of the situation in 2018. We do not assess the likely increase in vulnerabilities and deterioration of financial protection that has occurred since then, due to the onset of the COVID-19 pandemic in 2020 and the continued intensification of conflict violence, culminating in the Israel-Hamas war in 2023. These events have probably affected the OPT population in general, but even more acutely (with future long-term repercussions) in the Gaza strip. It is plausible to expect that the collapse of the Gazan health system in late 2023/early 2024 will lead to a stark increase in foregone healthcare, health-related financial catastrophe, and poverty. Negative reverberations at the health system level from the overall rise in violence may be expected also in the West Bank. Future research may apply our proposed methodology to provide a rounded assessment of these consequences in due course.

We note that implementation of continuous and enhanced data monitoring mechanisms will allow for real-time identification of shifts in health vulnerabilities [50]. This proactive approach ensures that future analyses go beyond static snapshots. At this point in time, the discussion in our study still provides insights about aspects – e.g., health financing and insurance bottlenecks, sub-groups particularly vulnerable to financial risk – that will deserve careful attention in future efforts to rebuild the Palestinian health system. These aspects will require addressing if the goal is to “build back better” the heath system, avoiding the presence of left-behind sub-populations with respect to access to quality care and protection from health-related financial risks [51, 52].

## Data Availability

All data produced are available online at https://www.pcbs.gov.ps/PCBS-Metadata-en-v5.2/index.php/catalog/629/data-dictionary/F3?file_name=sefsec%202018%2018+-E

## Acknowledgments

We thank Dr Abed Fadaleh (PA Ministry of Social Development), Dr Ahmed Qatou (PA Ministry of Health), Dr Rihab Quqa and Dr Zoheir Alkhatib (UNRWA-Gaza), Dr Samira Awawdeh, Dr Tareq Sadeq and Lama Shakhashi (Birzeit University), and former PA Deputy Minister of Social Development Dawoud Aldeek for feedback on this work. **Funding provided by the Centre for Health Economics, University of York, UK.**

## Footnotes

1 The concept of “de-development” in the Palestinian context refers to a process through which infrastructure and economic capacity have regressed or stifled, rather than simply underdeveloped.

2 https://www.unocha.org/publications/report/occupied-palestinian-territory/hostilities-gaza-strip-and-israel-flash-update-120

3 In coordination with the Food Security Sector (co-led by the FAO and WFP), supported by the United Nations Relief and Works Agency (UNRWA) and the Union of Agriculture Working Committee (UAWC).

4 We also use quintiles of the index to validate the results, see Appendix.

5 Details regarding these covariates can be found in the Appendix.

6 Appendix Table A3 shows incidence of CHE at various thresholds for both regions together and individually.

7 Coefficients for control variables are reported in the Appendix.

## Notes

### Competing Interest Statement

The authors have declared no competing interest.

### Funding Statement

This study was funded by the University of York Centre for Health Economics.

### Author Declarations

Palestinian Central Bureau of Statistics, Socio-Economic Conditions Survey 2018: https://www.pcbs.gov.ps/PCBS-Metadata-en-v5.2/index.php/catalog/629/data-dictionary/F3?file_name=sefsec%202018%2018+-E

