## Supplemental Appendix for "Multidimensional vulnerability and financial risk protection in health in contexts of protracted conflict: Evidence from the Occupied Palestinian Territory"

### SUPPLEMENTARY MATERIALS

#### .1. Variable Description.

- **NCDs and/or disability:** We indicate the presence of at least one HH member with Non-Communicable Diseases (NCDs) such as Diabetes mellitus, Hypertension, and Cardiovascular Diseases, and/or disability which includes difficulties in vision, hearing, movement, communication, focus, and memory. This variable is categorized into four groups:

-(Reference group): Households in which none of the members have NCDs or disabil- ity.

-Households with at least one member who has NCDs only.

-Households with at least one member who has disability only.

-Households with at least one member who has both NCDs and disabilities.

- **Employment status:** based on the household’s head employment status: not work- ing, working part-time and working full-time.
- **Household size:** A continuous variable indicating the number of HH members.
- **Insurance status:** This binary variable indicates whether the household head had insurance.
- **Insurance type:** indicates whether the household head has no health insurance at all (the reference group), has governmental health insurance only (PA only), has UNRWA health insurance only (UNRWA only), both governmental and UNRWA health insurance (PA + UNRWA) and others for other types of health insurance such as the private health insurance.
- **Educational level:** This variable is categorised into three groups indicating whether the level of education of the household head is less than secondary (the reference group), secondary or above secondary.

#### .2. Figures & Tables.

Table A1. CHE Mean differences By Region

### CHEs Consumption Exp. CHEs Nonfood Exp.

| CHEs | WB N=5875 | Gaza N=4018 | Mean Diff | p-value |  | WB N=5874 | Gaza N=4018 | Mean Diff | p-value |
| --- | --- | --- | --- | --- | --- | --- | --- | --- | --- |
| 5% | 0.413 | 0.363 | 0.05*** | 0.000 |  | 0.583 | 0.502 | 0.08*** | 0.000 |
| 10% | 0.191 | 0.171 | 0.02* | 0.011 |  | 0.352 | 0.274 | 0.08*** | 0.000 |
| 15% | 0.096 | 0.084 | 0.01* | 0.045 |  | 0.215 | 0.169 | 0.05*** | 0.000 |
| 20% | 0.057 | 0.046 | 0.01* | 0.017 |  | 0.134 | 0.103 | 0.03*** | 0.000 |
| 25% | 0.036 | 0.027 | 0.01* | 0.016 |  | 0.090 | 0.064 | 0.03*** | 0.000 |
| 30% | 0.024 | 0.017 | 0.01* | 0.022 |  | 0.062 | 0.044 | 0.02*** | 0.000 |
| 35% | 0.017 | 0.009 | 0.01*** | 0.001 |  | 0.043 | 0.029 | 0.01*** | 0.000 |
| 40% | 0.012 | 0.005 | 0.01*** | 0.001 |  | 0.032 | 0.020 | 0.01*** | 0.000 |

Table A2. Vulnerability Index Factor Loadings

WB	Gaza

|  | Loadings | Uniqueness | Loadings | Uniqueness |
| --- | --- | --- | --- | --- |
| Poverty Status (SA) | 0.666 | 0.557 | 0.685 | 0.530 |
| Financial Fragility (SA) | 0.397 | 0.843 | 0.541 | 0.707 |
| Need for Assistance (SA) | 0.739 | 0.455 | 0.733 | 0.463 |
| Other Shocks | 0.428 | 0.817 | 0.308 | 0.905 |
| Asset Ownership | 0.422 | 0.822 | 0.533 | 0.716 |
| Subjective Deprivation | 0.535 | 0.714 | 0.544 | 0.704 |
| Human Insecurity | 0.184 | 0.966 | 0.233 | 0.946 |

### Table A3. Incidence of Catastrophic Health Expenditure (CHEs)

**CHEs Consumption Exp.	CHEs Nonfood Exp.**

| CHEs at | All | WB | Gaza |  | All | WB | Gaza |
| --- | --- | --- | --- | --- | --- | --- | --- |
| 5% | 39.28 | 41.31 | 36.31 |  | 54.99 | 58.27 | 50.20 |
| 10% | 18.27 | 19.08 | 17.07 |  | 32.01 | 35.24 | 27.40 |
| 15% | 9.14 | 9.62 | 8.44 |  | 19.64 | 21.48 | 16.95 |
| 20% | 5.23 | 5.67 | 4.58 |  | 12.14 | 13.40 | 10.30 |
| 25% | 3.20 | 3.56 | 2.69 |  | 7.96 | 9.01 | 6.42 |
| 30% | 2.09 | 2.37 | 1.69 |  | 5.45 | 6.18 | 4.38 |
| 35% | 1.37 | 1.70 | 0.90 |  | 3.77 | 4.34 | 2.94 |
| 40% | 0.94 | 1.21 | 0.55 |  | 2.71 | 3.18 | 2.02 |

Table A4. Catastrophic Health Expenditure and Vulnerability (Terciles): Full Coefficients

**PANEL A: Consumption Expenditure vs. Non-food Expenditure**

| Dep Var: |  | CHE 10% |  |  | |  |  | Non-food 20% |  |
| --- | --- | --- | --- | --- | --- | --- | --- | --- | --- |
|  | (1) | (2) | (3) |  | | (4) | (5) |  | (6) |
| Odds Ratios | All | WB | Ghaza |  | | All | WB |  | Gaza |
| **index tercile = 2** | 1.264^∗∗^ | 1.135 | 1.500^∗∗^ | | 1.455^∗∗∗^ | | 1.420^∗∗^ | 1.552^∗∗∗^ | |
|  | (0.141) | (0.171) | (0.251) | | (0.141) | | (0.234) | (0.127) | |
| **index tercile = 3** | 1.762^∗∗∗^ | 1.846^∗∗∗^ | 1.617^∗∗∗^ | | 2.053^∗∗∗^ | | 2.085^∗∗∗^ | 1.920^∗∗∗^ | |
|  | (0.144) | (0.216) | (0.148) | | (0.241) | | (0.255) | (0.447) | |
| part time work | 0.752^∗∗^ | 0.690^∗∗∗^ | 0.810 | | 0.624^∗∗∗^ | | 0.549^∗∗∗^ | 0.727 | |
|  | (0.099) | (0.084) | (0.228) | | (0.078) | | (0.068) | (0.181) | |
| full time | 0.741^∗∗∗^ | 0.699^∗∗∗^ | 0.824 | | 0.627^∗∗∗^ | | 0.576^∗∗∗^ | 0.751^∗∗∗^ | |
|  | (0.071) | (0.092) | (0.105) | | (0.055) | | (0.065) | (0.083) | |
| preparatory | 0.901 | 0.850^∗∗^ | 1.045 | | 0.921 | | 0.917 | 1.006 | |
|  | (0.073) | (0.063) | (0.183) | | (0.098) | | (0.100) | (0.244) | |
| secondary | 0.774^∗∗∗^ | 0.743^∗∗∗^ | 0.867 | | 0.760^∗∗^ | | 0.704^∗∗^ | 0.913 | |
|  | (0.074) | (0.065) | (0.174) | | (0.083) | | (0.100) | (0.168) | |
| above secondary | 0.684^∗∗∗^ | 0.622^∗∗∗^ | 0.793 | | 0.711^∗∗∗^ | | 0.608^∗∗∗^ | 0.881 | |
|  | (0.074) | (0.053) | (0.179) | | (0.084) | | (0.054) | (0.220) | |
| NCDs only | 1.547^∗∗∗^ | 1.590^∗∗∗^ | 1.411^∗∗∗^ | | 1.761^∗∗∗^ | | 1.930^∗∗∗^ | 1.349^∗^ | |
|  | (0.102) | (0.138) | (0.141) | | (0.176) | | (0.153) | (0.222) | |
| Disability only | 2.306^∗∗∗^ | 2.342^∗∗∗^ | 2.183^∗∗∗^ | | 2.615^∗∗∗^ | | 2.617^∗∗∗^ | 2.408^∗∗∗^ | |
|  | (0.194) | (0.332) | (0.212) | | (0.338) | | (0.499) | (0.448) | |
| Both | 3.068^∗∗∗^ | 3.355^∗∗∗^ | 2.620^∗∗∗^ | | 3.315^∗∗∗^ | | 3.857^∗∗∗^ | 2.521^∗∗∗^ | |
|  | (0.246) | (0.298) | (0.357) | | (0.327) | | (0.365) | (0.391) | |
| PA only | 1.489^∗∗∗^ | 1.354^∗∗∗^ | 2.528^∗∗∗^ | | 1.469^∗∗∗^ | | 1.365^∗∗∗^ | 2.002^∗∗∗^ | |
|  | (0.126) | (0.089) | (0.632) | | (0.111) | | (0.122) | (0.364) | |
| UNRWA only | 0.975 | 0.962 | 1.450 | | 0.930 | | 0.968 | 1.041 | |
|  | (0.124) | (0.147) | (0.471) | | (0.121) | | (0.155) | (0.308) | |
| PA+UNRWA | 1.209 | 1.008 | 2.032^∗^ | | 1.109 | | 0.949 | 1.532 | |
|  | (0.214) | (0.188) | (0.767) | | (0.166) | | (0.220) | (0.420) | |
| others | 0.910 | 0.846 | 1.590 | | 0.825 | | 0.764 | 1.545 | |
|  | (0.282) | (0.293) | (1.438) | | (0.295) | | (0.314) | (1.724) | |
| HH size | 0.874^∗∗∗^ | 0.872^∗∗∗^ | 0.880^∗∗∗^ | | 0.856^∗∗∗^ | | 0.851^∗∗∗^ | 0.869^∗∗∗^ | |
|  | (0.013) | (0.022) | (0.013) | | (0.020) | | (0.033) | (0.025) | |
| Governorate FE | Yes | Yes | Yes | | Yes | | Yes | Yes | |
| *Observations* | 9647 | 5801 | 3846 | | 9646 | | 5800 | 3846 | |
| *Clusters − Governorate* | 16 | 11 | 5 | | 16 | | 11 | 5 | |

Exponentiated coefficients; Standard errors in parentheses

SE clustered at governorate level. Governorate fixed effects in all models. All control variable coefficients reported in Appendix

^∗^ *p <* 0*.*10, ^∗∗^ *p <* 0*.*05, ^∗∗∗^ *p <* 0*.*01

width=12.8cm

Table A5. Catastrophic Health Expenditures (Terciles), Alternate Specifications

(1)	(2)	(3)	(4)	(5)	(6)

Dep: Var: CHE-10%	All	WB	Gaza	All	WB	Gaza

comp idx terc=2	1.327^∗∗^	1.185	1.579^∗∗^	1.296^∗∗^	1.157	1.549^∗∗∗^

(0.149)	(0.171)	(0.283)	(0.137)	(0.162)	(0.243)

comp idx terc=3	1.958^∗∗∗^ 2.059^∗∗∗^ 1.768^∗∗∗^ 1.891^∗∗∗^ 1.973^∗∗∗^ 1.709^∗∗∗^

(0.154)	(0.247)	(0.157)	(0.138)	(0.215)	(0.119)

part time	0.668^∗∗∗^ 0.650^∗∗∗^	0.691

(0.091)	(0.078)	(0.205)

full time	0.726^∗∗∗^ 0.695^∗∗∗^	0.824

(0.081)	(0.098)	(0.123)

long working hours	0.693^∗∗∗^ 0.692^∗∗∗^	0.729^∗^

(0.060)	(0.063)	(0.127)

preparatory	0.809^∗∗^	0.787^∗∗^	0.891	0.794^∗∗^	0.779^∗∗^	0.875

(0.073)	(0.092)	(0.133)	(0.072)	(0.090)	(0.134)

secondary	0.712^∗∗∗^ 0.674^∗∗∗^	0.805	0.698^∗∗∗^ 0.664^∗∗∗^	0.795

(0.073)	(0.090)	(0.122)	(0.067)	(0.079)	(0.122)

above secondary	0.710^∗∗∗^ 0.650^∗∗∗^	0.804^∗^	0.686^∗∗∗^ 0.630^∗∗∗^	0.796^∗∗^

(0.049)	(0.051)	(0.094)	(0.050)	(0.055)	(0.092)

chronic only	1.530^∗∗∗^ 1.583^∗∗∗^ 1.410^∗∗∗^ 1.547^∗∗∗^ 1.598^∗∗∗^ 1.390^∗∗∗^

(0.104)	(0.123)	(0.147)	(0.102)	(0.134)	(0.137)

disability only	1.779^∗∗∗^ 1.840^∗∗∗^ 1.717^∗∗∗^ 1.769^∗∗∗^ 1.808^∗∗∗^ 1.679^∗∗∗^

(0.196)	(0.196)	(0.335)	(0.187)	(0.175)	(0.319)

chronic and disability	2.771^∗∗∗^ 3.328^∗∗∗^ 2.128^∗∗∗^ 2.834^∗∗∗^ 3.410^∗∗∗^ 2.158^∗∗∗^

(0.322)	(0.464)	(0.313)	(0.300)	(0.470)	(0.236)

Official refugee status	0.806^∗∗^	0.750^∗∗^	0.859

(0.084)	(0.084)	(0.168)

HH size	0.904^∗∗∗^ 0.902^∗∗∗^ 0.910^∗∗∗^ 0.913^∗∗∗^ 0.911^∗∗∗^ 0.921^∗∗∗^ (0.011)	(0.019)	(0.015)	(0.010)	(0.016)	(0.013)

one working HH member	0.712^∗∗∗^ 0.641^∗∗∗^	0.798

(0.063)	(0.043)	(0.123)

at least 2 working	0.631^∗∗∗^ 0.634^∗∗∗^ 0.487^∗∗∗^ (0.064)	(0.038)	(0.131)

PA only	1.432^∗∗∗^ 1.283^∗∗∗^ 2.568^∗∗∗^

(0.158)	(0.103)	(0.819)

UNRWA only	0.994	0.878	1.763^∗^

(0.123)	(0.118)	(0.590)

PA+UNRWA	1.133	1.008	1.997

(0.215)	(0.206)	(0.879)

others	0.911	0.779	2.688

(0.250)	(0.222)	(1.830)

Governorate FE	Yes	Yes	Yes	Yes	Yes	Yes

*Observations*	9642	5799	3843	9642	5799	3843

*Clusters − Governorate*	16	11	5	16	11	5

Exponentiated coefficients; Standard errors in parentheses SE clustered at governorate level

^∗^ *p <* 0*.*10, ^∗∗^ *p <* 0*.*05, ^∗∗∗^ *p <* 0*.*01

width=12.6cm

Table A6. Catastrophic Health Expenditures (Quantiles), Alternate Specifications

(1)	(2)	(3)	(4)	(5)	(6)

Dep: Var: CHE-10%	All	WB	Gaza	All	WB	Gaza

comp idx 5q=2	1.472^∗∗∗^	1.355	1.659^∗∗^ 1.466^∗∗∗^ 1.358^∗^	1.649^∗∗^ (0.218) (0.278) (0.405) (0.204) (0.252) (0.409)

comp idx 5q=3	1.623^∗∗∗^	1.399	2.032^∗∗∗^ 1.596^∗∗∗^	1.386	1.995^∗∗∗^ (0.213) (0.310) (0.269) (0.203) (0.305) (0.223)

comp idx 5q=4	1.873^∗∗∗^ 1.942^∗∗∗^ 1.730^∗∗∗^ 1.829^∗∗∗^ 1.900^∗∗∗^ 1.682^∗∗∗^ (0.140) (0.215) (0.195) (0.129) (0.191) (0.187)

comp idx 5q=5	2.580^∗∗∗^ 2.704^∗∗∗^ 2.342^∗∗∗^ 2.490^∗∗∗^ 2.602^∗∗∗^ 2.246^∗∗∗^ (0.239) (0.424) (0.151) (0.218) (0.378) (0.157)

part time	0.671^∗∗∗^ 0.652^∗∗∗^	0.695

(0.091) (0.077) (0.208)

full time	0.730^∗∗∗^ 0.703^∗∗^	0.824

(0.081) (0.099) (0.126)

long working hours	0.698^∗∗∗^ 0.703^∗∗∗^ 0.732^∗^ (0.063) (0.066) (0.129)

preparatory	0.810^∗∗^	0.785^∗^	0.894	0.796^∗∗^ 0.779^∗∗^	0.879

(0.076) (0.101) (0.124) (0.075) (0.098) (0.127)

secondary	0.722^∗∗∗^ 0.679^∗∗∗^	0.823	0.709^∗∗∗^ 0.670^∗∗∗^	0.813

(0.074) (0.093) (0.117) (0.068) (0.083) (0.116)

above secondary	0.726^∗∗∗^ 0.664^∗∗∗^	0.830	0.702^∗∗∗^ 0.645^∗∗∗^ 0.821^∗^ (0.053) (0.057) (0.097) (0.054) (0.059) (0.095)

chronic only	1.540^∗∗∗^ 1.594^∗∗∗^ 1.404^∗∗∗^ 1.557^∗∗∗^ 1.605^∗∗∗^ 1.385^∗∗∗^ (0.106) (0.119) (0.154) (0.103) (0.129) (0.146)

disability only	1.761^∗∗∗^ 1.814^∗∗∗^ 1.707^∗∗∗^ 1.752^∗∗∗^ 1.783^∗∗∗^ 1.672^∗∗∗^ (0.195) (0.201) (0.337) (0.185) (0.177) (0.320)

chronic and disability	2.760^∗∗∗^ 3.312^∗∗∗^ 2.134^∗∗∗^ 2.822^∗∗∗^ 3.386^∗∗∗^ 2.165^∗∗∗^ (0.318) (0.467) (0.309) (0.290) (0.458) (0.229)

Official refugee status	0.807^∗∗^ 0.756^∗∗^	0.855

(0.084) (0.083) (0.164)

HH size	0.904^∗∗∗^ 0.902^∗∗∗^ 0.910^∗∗∗^ 0.913^∗∗∗^ 0.911^∗∗∗^ 0.921^∗∗∗^ (0.011) (0.018) (0.014) (0.009) (0.015) (0.013)

one working member in the household	0.716^∗∗∗^ 0.647^∗∗∗^	0.799

(0.064) (0.043) (0.124)

at least 2 working members in the household	0.641^∗∗∗^ 0.646^∗∗∗^ 0.494^∗∗∗^ (0.066) (0.036) (0.134)

PA only	1.426^∗∗∗^ 1.271^∗∗∗^ 2.561^∗∗∗^

(0.157) (0.101) (0.791)

UNRWA only	0.991	0.876	1.751^∗^

(0.122) (0.116) (0.587)

PA+UNRWA	1.124	1.009	1.975

(0.210) (0.200) (0.857)

others	0.910	0.782	2.819

(0.246) (0.218) (1.992)

Governorate FE	Yes	Yes	Yes	Yes	Yes	Yes

*Observations*	9641	5798	3843	9641	5798	3843

*Clusters − Governorate*	16	11	5	16	11	5

Exponentiated coefficients; Standard errors in parentheses SE clustered at governorate level

^∗^ *p <* 0*.*10, ^∗∗^ *p <* 0*.*05, ^∗∗∗^ *p <* 0*.*01

width=13.7cm

Table A7. Catex 10%: MPCE vs Vulnerability Index (OR): All coefficients

Quintile Definition:	**MCPE	Vulnerability Index**

|  | (1) | (2) | (3) | (4) | (5) | (6) |
| --- | --- | --- | --- | --- | --- | --- |
| Dep: Var: CHE-10% | All | WB | Gaza | All | WB | Gaza |
| **Quintle=2** | 0.917 | 0.834^∗^ | 1.088 | 1.448^∗∗∗^ | 1.447^∗∗^ | 1.463^∗^ |
|  | (0.089) | (0.086) | (0.173) | (0.181) | (0.272) | (0.289) |
| **Quintle=3** | 0.958 | 0.931 | 1.058 | 1.588^∗∗∗^ | 1.484 | 1.782^∗∗∗^ |
|  | (0.095) | (0.101) | (0.204) | (0.218) | (0.379) | (0.201) |
| **Quintle=4** | 1.068 | 0.893 | 1.515^∗^ | 1.740^∗∗∗^ | 1.851^∗∗∗^ | 1.571^∗∗∗^ |
|  | (0.143) | (0.118) | (0.338) | (0.126) | (0.214) | (0.125) |
| **Quintle=5** | 0.994 | 0.836 | 1.499 | 2.280^∗∗∗^ | 2.486^∗∗∗^ | 1.963^∗∗∗^ |
|  | (0.160) | (0.115) | (0.462) | (0.232) | (0.398) | (0.161) |
| part time | 0.745^∗∗^ | 0.686^∗∗∗^ | 0.813 | 0.756^∗∗^ | 0.690^∗∗∗^ | 0.811 |
|  | (0.101) | (0.087) | (0.226) | (0.099) | (0.083) | (0.226) |
| full time | 0.679^∗∗∗^ | 0.636^∗∗∗^ | 0.775^∗∗^ | 0.746^∗∗∗^ | 0.705^∗∗∗^ | 0.825 |
|  | (0.063) | (0.079) | (0.094) | (0.070) | (0.093) | (0.102) |
| preparatory | 0.857^∗∗^ | 0.815^∗∗∗^ | 0.987 | 0.903 | 0.849^∗∗^ | 1.046 |
|  | (0.060) | (0.060) | (0.136) | (0.074) | (0.067) | (0.175) |
| secondary | 0.712^∗∗∗^ | 0.681^∗∗∗^ | 0.787^∗^ | 0.788^∗∗^ | 0.753^∗∗∗^ | 0.879 |
|  | (0.060) | (0.071) | (0.101) | (0.077) | (0.070) | (0.166) |
| above secondary | 0.587^∗∗∗^ | 0.543^∗∗∗^ | 0.631^∗∗∗^ | 0.707^∗∗∗^ | 0.643^∗∗∗^ | 0.808 |
|  | (0.061) | (0.062) | (0.110) | (0.076) | (0.061) | (0.174) |
| NCDs only | 1.508^∗∗∗^ | 1.597^∗∗∗^ | 1.300^∗^ | 1.568^∗∗∗^ | 1.616^∗∗∗^ | 1.414^∗∗∗^ |
|  | (0.105) | (0.121) | (0.188) | (0.102) | (0.135) | (0.140) |
| Disability only | 2.504^∗∗∗^ | 2.614^∗∗∗^ | 2.248^∗∗∗^ | 2.297^∗∗∗^ | 2.341^∗∗∗^ | 2.165^∗∗∗^ |
|  | (0.237) | (0.445) | (0.224) | (0.199) | (0.338) | (0.233) |
| Both | 3.149^∗∗∗^ | 3.655^∗∗∗^ | 2.455^∗∗∗^ | 3.064^∗∗∗^ | 3.346^∗∗∗^ | 2.611^∗∗∗^ |
|  | (0.292) | (0.387) | (0.356) | (0.251) | (0.310) | (0.357) |
| PA only | 1.507^∗∗∗^ | 1.369^∗∗∗^ | 2.267^∗∗∗^ | 1.491^∗∗∗^ | 1.353^∗∗∗^ | 2.527^∗∗∗^ |
|  | (0.129) | (0.094) | (0.563) | (0.126) | (0.087) | (0.629) |
| UNRWA only | 1.045 | 1.050 | 1.335 | 0.978 | 0.965 | 1.443 |
|  | (0.128) | (0.147) | (0.457) | (0.125) | (0.148) | (0.480) |
| PA+UNRWA | 1.246 | 1.098 | 1.821 | 1.203 | 1.014 | 2.009^∗^ |
|  | (0.207) | (0.217) | (0.665) | (0.211) | (0.186) | (0.758) |
| others | 0.903 | 0.882 | 1.043 | 0.907 | 0.846 | 1.661 |
|  | (0.279) | (0.303) | (1.008) | (0.281) | (0.290) | (1.550) |
| HH size | 0.886^∗∗∗^ | 0.873^∗∗∗^ | 0.924^∗∗∗^ | 0.874^∗∗∗^ | 0.874^∗∗∗^ | 0.880^∗∗∗^ |
|  | (0.014) | (0.021) | (0.017) | (0.012) | (0.022) | (0.012) |
| Governorate FE | Yes | Yes | Yes | Yes | Yes | Yes |
| *Observations* | 9887 | 5872 | 4015 | 9647 | 5801 | 3846 |
| *Clusters − Governorate* | 16 | 11 | 5 | 16 | 11 | 5 |

Exponentiated coefficients; Standard errors in parentheses SE clustered at governorate level

^∗^ *p <* 0*.*10, ^∗∗^ *p <* 0*.*05, ^∗∗∗^ *p <* 0*.*01

Table A8. Catastrophic Health Expenditures, By Insurance Type: all coefficients

Insurance Type:	**PA Only	UNRWA Only**

|  | (1) | (2) | (3) | (4) | (5) | (6) |
| --- | --- | --- | --- | --- | --- | --- |
| Dep: Var: CHE-10% | All | WB | Gaza | All | WB | Gaza |
| **index tercile=2** | 1.331^∗∗^ | 1.133 | 1.876^∗∗∗^ | 1.122 | 1.225 | 1.109 |
|  | (0.178) | (0.153) | (0.398) | (0.229) | (0.343) | (0.409) |
| **index tercile=3** | 1.787^∗∗∗^ | 1.877^∗∗∗^ | 1.583^∗∗∗^ | 1.523 | 1.481 | 1.623 |
|  | (0.172) | (0.250) | (0.219) | (0.402) | (0.519) | (0.752) |
| part time | 0.635^∗∗^ | 0.679 | 0.557^∗∗∗^ | 0.379^∗∗∗^ | 0.250^∗∗∗^ | 0.565 |
|  | (0.113) | (0.164) | (0.120) | (0.104) | (0.079) | (0.208) |
| full time | 0.656^∗∗∗^ | 0.670^∗∗^ | 0.657^∗^ | 0.661 | 0.594 | 0.730 |
|  | (0.085) | (0.123) | (0.159) | (0.172) | (0.247) | (0.271) |
| long working hours | 0.719^∗∗∗^ | 0.713^∗∗^ | 0.783 | 0.452^∗∗∗^ | 0.453^∗∗^ | 0.420^∗∗∗^ |
|  | (0.092) | (0.098) | (0.251) | (0.067) | (0.139) | (0.021) |
| preparatory | 0.792^∗∗^ | 0.815 | 0.778 | 0.731 | 0.524 | 1.093 |
|  | (0.091) | (0.129) | (0.121) | (0.218) | (0.241) | (0.345) |
| secondary | 0.757^∗∗^ | 0.685^∗∗∗^ | 0.923 | 0.534^∗∗^ | 0.493^∗^ | 0.604 |
|  | (0.087) | (0.073) | (0.198) | (0.130) | (0.201) | (0.198) |
| above secondary | 0.650^∗∗∗^ | 0.632^∗∗∗^ | 0.726 | 0.723 | 0.635 | 0.906 |
|  | (0.082) | (0.090) | (0.196) | (0.143) | (0.180) | (0.203) |
| chronic only | 1.833^∗∗∗^ | 1.745^∗∗∗^ | 2.091^∗∗∗^ | 1.464 | 1.266 | 1.867^∗∗^ |
|  | (0.166) | (0.184) | (0.445) | (0.365) | (0.491) | (0.585) |
| disability only | 1.813^∗∗∗^ | 1.753^∗∗∗^ | 1.873^∗∗∗^ | 1.516 | 1.664 | 1.418 |
|  | (0.258) | (0.367) | (0.430) | (0.440) | (0.790) | (0.595) |
| chronic and disability | 3.364^∗∗∗^ | 3.708^∗∗∗^ | 2.520^∗∗∗^ | 2.527^∗∗∗^ | 2.451^∗∗∗^ | 2.502^∗∗∗^ |
|  | (0.527) | (0.769) | (0.581) | (0.514) | (0.723) | (0.586) |
| HH size | 0.932^∗∗∗^ | 0.930^∗∗^ | 0.939^∗∗^ | 0.880^∗∗∗^ | 0.902 | 0.865^∗∗∗^ |
|  | (0.020) | (0.031) | (0.028) | (0.036) | (0.076) | (0.021) |
| Governorate FE | Yes | Yes | Yes | Yes | Yes | Yes |
| *Observations* | 3246 | 2212 | 1034 | 1352 | 728 | 624 |
| *Clusters − Governorate* | 16 | 11 | 5 | 16 | 11 | 5 |

Exponentiated coefficients; Standard errors in parentheses SE clustered at governorate level

^∗^ *p <* 0*.*10, ^∗∗^ *p <* 0*.*05, ^∗∗∗^ *p <* 0*.*01

Table A9. Catastrophic Health Expenditures, By Insurance Status: all coefficients

| Insurance Status: |  | **Insured** |  |  | **Uninsured** |  |
| --- | --- | --- | --- | --- | --- | --- |
|  | (1) | (2) | (3) | (4) | (5) | (6) |
| Dep: Var: CHE-10% | All | WB | Gaza | All | WB | Gaza |

| **index tercile=2** | 1.334^∗∗^ | 1.121 | 1.620^∗∗∗^ | 1.337^∗^ | 1.337 | 1.312 |
| --- | --- | --- | --- | --- | --- | --- |
|  | (0.172) | (0.168) | (0.300) | (0.226) | (0.236) | (1.010) |
| **index tercile=3** | 1.825^∗∗∗^ | 1.820^∗∗∗^ | 1.767^∗∗∗^ | 2.622^∗∗∗^ | 2.571^∗∗∗^ | 2.339 |
|  | (0.172) | (0.307) | (0.157) | (0.356) | (0.343) | (2.540) |
| part time | 0.638^∗∗∗^ | 0.633^∗∗∗^ | 0.644 | 0.815 | 0.693^∗^ | 2.907 |
|  | (0.099) | (0.089) | (0.195) | (0.167) | (0.132) | (2.418) |
| full time | 0.730^∗∗∗^ | 0.722^∗^ | 0.773 | 0.798 | 0.690^∗∗∗^ | 3.116^∗∗^ |
|  | (0.089) | (0.134) | (0.124) | (0.114) | (0.092) | (1.784) |
| long working hours | 0.674^∗∗∗^ | 0.665^∗∗∗^ | 0.717^∗∗^ | 0.850 | 0.770 | 1.322 |
|  | (0.072) | (0.104) | (0.116) | (0.141) | (0.131) | (1.298) |
| preparatory | 0.835^∗^ | 0.837 | 0.875 | 0.664^∗∗∗^ | 0.653^∗∗∗^ | 1.050 |
|  | (0.086) | (0.128) | (0.128) | (0.084) | (0.088) | (0.595) |
| secondary | 0.704^∗∗∗^ | 0.672^∗∗∗^ | 0.771^∗^ | 0.641^∗∗^ | 0.623^∗∗^ | 0.700 |
|  | (0.072) | (0.087) | (0.116) | (0.112) | (0.116) | (0.503) |
| above secondary | 0.713^∗∗∗^ | 0.635^∗∗∗^ | 0.814 | 0.533^∗∗∗^ | 0.566^∗∗^ | 0.261^∗^ |
|  | (0.063) | (0.076) | (0.105) | (0.119) | (0.139) | (0.212) |
| chronic only | 1.414^∗∗∗^ | 1.496^∗∗∗^ | 1.317^∗∗∗^ | 1.934^∗∗∗^ | 1.770^∗∗∗^ | 5.743^∗∗^ |
|  | (0.110) | (0.155) | (0.120) | (0.309) | (0.274) | (4.486) |
| disability only | 1.689^∗∗∗^ | 1.821^∗∗∗^ | 1.587^∗∗^ | 1.890^∗∗^ | 1.631^∗^ | 5.204^∗∗^ |
|  | (0.200) | (0.217) | (0.301) | (0.547) | (0.476) | (3.871) |
| chronic and disability | 2.582^∗∗∗^ | 3.204^∗∗∗^ | 1.996^∗∗∗^ | 3.221^∗∗∗^ | 3.459^∗∗∗^ | 2.034 |
|  | (0.304) | (0.459) | (0.303) | (0.778) | (0.778) | (3.718) |
| HH size | 0.902^∗∗∗^ | 0.903^∗∗∗^ | 0.906^∗∗∗^ | 0.897^∗∗∗^ | 0.894^∗∗∗^ | 0.909 |
|  | (0.012) | (0.021) | (0.016) | (0.024) | (0.025) | (0.110) |
| Governorate FE | Yes | Yes | Yes | Yes | Yes | Yes |
| *Observations* | 7670 | 4063 | 3607 | 1972 | 1736 | 236 |
| *Clusters − Governorate* | 16 | 11 | 5 | 16 | 11 | 5 |

Exponentiated coefficients; Standard errors in parentheses SE clustered at governorate level

^∗^ *p <* 0*.*10, ^∗∗^ *p <* 0*.*05, ^∗∗∗^ *p <* 0*.*01

| Insurance Status: |  | **Urban** |  | **Rural** | **Camps** |
| --- | --- | --- | --- | --- | --- |
|  | (1) | (2) | (3) | (4) | (5) |
| Dep: Var: CHE-10% | All | WB | Gaza | All | All |
| Index Tercile =2 | 1.403^∗∗∗^ | 1.212 | 1.740^∗∗∗^ | 0.987 | 1.115 |
| Index Tercile =3 | (0.157)  1.966^∗∗∗^ | (0.161)  1.979^∗∗∗^ | (0.271)  1.987^∗∗∗^ | (0.245) 2.077^∗∗^ | (0.179) 1.648^∗∗^ |
|  | (0.112) | (0.184) | (0.136) | (0.591) | (0.384) |
| part time | 0.705^∗∗^ | 0.664^∗∗∗^ | 0.737 | 0.718^∗^ | 0.391^∗∗∗^ |
|  | (0.113) | (0.080) | (0.248) | (0.133) | (0.131) |
| full time | 0.735^∗∗∗^ | 0.728^∗∗^ | 0.756^∗∗^ | 0.586^∗∗^ | 0.965 |
|  | (0.083) | (0.118) | (0.107) | (0.131) | (0.227) |
| long working hours | 0.695^∗∗∗^ | 0.709^∗∗∗^ | 0.688^∗^ | 0.609^∗^ | 1.125 |
|  | (0.065) | (0.067) | (0.137) | (0.165) | (0.248) |
| preparatory | 0.867 | 0.818 | 0.973 | 0.729^∗∗∗^ | 0.481^∗∗∗^ |
|  | (0.097) | (0.124) | (0.166) | (0.064) | (0.076) |
| secondary | 0.738^∗∗∗^ | 0.685^∗∗^ | 0.841 | 0.599^∗∗∗^ | 0.646^∗∗^ |
|  | (0.084) | (0.109) | (0.131) | (0.086) | (0.114) |
| above secondary | 0.723^∗∗∗^ | 0.629^∗∗∗^ | 0.876 | 0.671 | 0.511^∗∗^ |
|  | (0.063) | (0.090) | (0.082) | (0.174) | (0.150) |
| chronic only | 1.535^∗∗∗^ | 1.537^∗∗∗^ | 1.525^∗∗∗^ | 1.294 | 1.236 |
| disability only | (0.157)  1.624^∗∗∗^ | (0.215)  1.659^∗∗∗^ | (0.229) 1.564^∗^ | (0.291)  2.234^∗∗∗^ | (0.470)  1.971^∗∗∗^ |
| chronic and disability | (0.247)  2.762^∗∗∗^ | (0.264)  3.402^∗∗∗^ | (0.378)  2.197^∗∗∗^ | (0.461)  2.389^∗∗∗^ | (0.456) 1.807^∗^ |
| HH size | (0.328)  0.904^∗∗∗^ | (0.535)  0.895^∗∗∗^ | (0.286)  0.914^∗∗∗^ | (0.662)  0.916^∗∗∗^ | (0.621)  0.821^∗∗∗^ |
|  | (0.014) | (0.026) | (0.020) | (0.023) | (0.048) |
| PA only | 1.408^∗∗^ | 1.243 | 2.576^∗∗∗^ | 1.449^∗∗∗^ | 1.085 |
|  | (0.213) | (0.168) | (0.856) | (0.187) | (0.586) |
| UNRWA only | 0.907 | 0.819 | 1.593 | 1.207 | 1.276 |
|  | (0.153) | (0.187) | (0.622) | (0.214) | (0.599) |
| PA+UNRWA | 1.025 | 0.888 | 1.824 | 1.126 | 1.217 |
|  | (0.215) | (0.220) | (0.830) | (0.191) | (0.552) |
| others | 0.828 | 0.679 | 2.839 | 1.492 | 0.201 |
|  | (0.288) | (0.232) | (2.241) | (0.526) | (0.257) |
| Governorate FE | Yes | Yes | Yes | Yes | Yes |
| *Observations* | 7253 | 4008 | 3245 | 1417 | 972 |
| *Clusters − Governorate* | 16 | 11 | 5 | 11 | 14 |

Exponentiated coefficients; Standard errors in parentheses SE clustered at governorate level

^∗^ *p <* 0*.*10, ^∗∗^ *p <* 0*.*05, ^∗∗∗^ *p <* 0*.*01

width=10.1cm

Table A11. Catastrophic Health Expenditures (Quantiles) Alternative com-

posite index: full coefficients

| Dep: Var: CHE-10% | (1) | (2) | (3) |
| --- | --- | --- | --- |
| Odd-ratios | All | WB | Gaza |
| Index Quintile New=2 | 1.548^∗∗∗^ | 1.548^∗∗∗^ | 1.526^∗∗∗^ |
| Index Quintile New=3 | (0.159)  1.667^∗∗∗^ | (0.239)  1.724^∗∗∗^ | (0.187)  1.589^∗∗∗^ |
| Index Quintile New=4 | (0.103)  1.801^∗∗∗^ | (0.141)  2.072^∗∗∗^ | (0.186)  1.448^∗∗∗^ |
| Index Quintile New=5 | (0.172)  2.535^∗∗∗^ | (0.224)  2.789^∗∗∗^ | (0.207)  2.128^∗∗∗^ |
|  | (0.227) | (0.370) | (0.185) |
| part time | 0.715^∗∗∗^ | 0.672^∗∗∗^ | 0.748 |
| full time | (0.083)  0.719^∗∗∗^ | (0.069)  0.690^∗∗∗^ | (0.181)  0.764^∗^ |
|  | (0.064) | (0.065) | (0.119) |
| long working hour | 0.812^∗∗^ | 0.771^∗∗∗^ | 0.899 |
|  | (0.070) | (0.063) | (0.159) |
| preparatory | 0.795^∗∗∗^ | 0.743^∗∗∗^ | 0.922 |
| secondary | (0.064)  0.696^∗∗∗^ | (0.079)  0.639^∗∗∗^ | (0.105)  0.815^∗∗∗^ |
|  | (0.057) | (0.074) | (0.053) |
| above secondary | 0.753^∗∗∗^ | 0.688^∗∗∗^ | 0.872 |
| chronic only | (0.065)  1.553^∗∗∗^ | (0.072)  1.568^∗∗∗^ | (0.111)  1.451^∗∗∗^ |
| disability only | (0.086)  1.772^∗∗∗^ | (0.133)  1.734^∗∗∗^ | (0.048)  1.789^∗∗∗^ |
| chronic and disability | (0.184)  2.721^∗∗∗^ | (0.256)  3.158^∗∗∗^ | (0.295)  2.258^∗∗∗^ |
| PA only | (0.223)  1.347^∗∗∗^ | (0.396)  1.234^∗∗∗^ | (0.178)  1.913^∗∗∗^ |
|  | (0.091) | (0.069) | (0.301) |
| UNRWA only | 0.966 | 1.010 | 1.202 |
|  | (0.085) | (0.110) | (0.239) |
| PA+UNRWA | 1.143 | 1.151 | 1.483 |
|  | (0.149) | (0.187) | (0.380) |
| others | 0.959 | 0.907 | 1.724 |
|  | (0.231) | (0.239) | (1.060) |
| Rural | 1.141 | 1.099 |  |
|  | (0.180) | (0.186) |  |
| Camps | 1.012 | 0.664 | 1.279 |
| HH size | (0.193)  0.903^∗∗∗^ | (0.175)  0.907^∗∗∗^ | (0.271)  0.904^∗∗∗^ |
|  | (0.008) | (0.015) | (0.007) |
| received any assistance | 1.083 | 1.123 | 1.103 |
|  | (0.073) | (0.172) | (0.097) |
| Governorate FE | Yes | Yes | Yes |
| *Observations* | 9912 | 5889 | 4023 |
| *Clusters − Governorate* | 16 | 11 | 5 |
